# Prefrontal-Limbic Dysconnectivity Underlies Emotion Regulation Deficits in Depressed Adolescents with Anxiety Disorder

**DOI:** 10.1101/2025.10.06.25337160

**Authors:** Nan Qiu, Shaoqing Li, Jiang Wu, Benjamin Becker, Wanhong Peng, Kena Li, Guocheng Zhao, Lan Hu, Hongmei Yan, Dezhong Yao

## Abstract

**Objective:** Major depressive disorder (MDD) in adolescence is frequently accompanied by emotion regulation (ER) deficits, particularly in socially stressful contexts. However, how comorbid social anxiety disorder (SAD) modulates ER during social exclusion in depressed adolescents the behavioral and neural level remains poorly understood.

**Method:** Sixty-two adolescents (30 with MDD, 32 healthy controls; aged 12–18) completed a social-exclusion-stimuli task during electroencephalogram (EEG) recording. Using EEG in combination with a dedicated paradigm showing social exclusion (versus neutral situations), ER ability was assessed via cognitive reappraisal performance, negative affect ratings, EEG and prefrontal–limbic functional connectivity. The Adolescent-MDD group was further stratified by SAD comorbidity status.

**Results:** Compared to healthy peers, adolescents with MDD showed reduced cognitive reappraisal use and heightened negative affect during social exclusion, reflecting impaired top-down control. Neurally, they showed increased LPP amplitudes and greater left prefrontal theta-band activity during reappraisal, alongside weakened prefrontal-limbic functional coupling. Critically, youth with comorbid SAD demonstrated more pronounced ER impairments and fronto-limbic dysconnectivity, suggesting SAD amplifies neural inefficiency and social withdrawal in depression.

**Conclusion:** Disrupted prefrontal–limbic connectivity may underlie ER impairments in depressed adolescents, particularly those with comorbid SAD. These findings identify a potential neurophysiological target for early intervention and offer mechanistic insight into the interaction of depression, social anxiety, and ER during adolescence.

## Introduction

Adolescence is a pivotal phase of transition from childhood to adulthood, marked by the fast maturation of the brain and substantial changes in social interactions.^1,2^ During this stage, the adolescent brain undergoes critical maturational stages while the risk for mental disorders including depression^3^ and external stressors, such as social interaction problems, academic pressure, peer competition, increase leading to substantial stress and the need for adaptive emotional regulation (ER).^4,5^

The capacity to adaptively regulate emotional states is essential for mental health, and dysfunctions in this domain contribute to the development of emotional disorders such as depression. Research indicates that adolescence is a high-risk period for the development of depression^6^ with overlapping and distinct symptoms and neurobiological signatures as adult depression.^3^ The ability to effectively regulate negative emotional experiences is a crucial factor in preventing depression.^7^ Studies indicate that adolescents with depression commonly exhibit ER deficits, manifested by diminished use of cognitive reappraisal, prolonged unpleasant emotional states, and lowered thresholds for emotional responses.^8^ Conversely, emotion regulation deficiencies worsen depressive symptoms, impair social functioning including interpersonal skills or academic performance, and increase the likelihood of comorbidities like anxiety disorders.^9^ Neuroimaging studies have identified abnormal activity patterns in the prefrontal-limbic network in depressed adolescents.^10^ Specifically, during emotion regulation tasks, dorsolateral prefrontal cortex (DLPFC) activation levels were significantly lower than those in healthy controls (HCs), whereas amygdala reactivity was excessively increased.^11^ The dysfunction in “top-down” regulation is believed as the core neural basis of emotional dysregulation and has been observed across a range of mental disorders characterized by emotion regulation impairments (e.g. Zimmermann et al. 2017).^12^

Social exclusion, a common negative event in adolescence, poses a particular challenge to adaptive emotion regulation in adolescence given their higher sensitivity of social exclusion, which is rooted in the rapid development of their social cognitive network, including increased responsivity of regions such as the anterior cingulate cortex (ACC) and medial prefrontal cortex (mPFC) to social feedback.^13^ Previous findings indicated that depressed adolescents exhibited stronger negative emotional responses to virtual social exclusion than healthy peers. For example, significant differences were observed for their autonomic nervous system activity indexed by lower heart rate variability and stress hormone levels in the context of higher cortisol levels^14^. Longitudinal studies further corroborate that teenagers who frequently endure social exclusion are 2-3 times more likely to acquire depressive disorders in early adulthood (see also Liu et al. 2025),^2,15^ emphasizing the pivotal role of social context in the development of emotional pathology during adolescence.^16^

Social anxiety disorder (SAD) frequently emerges during adolescence and exhibits a high comorbidity with depression. SAD is characterized by pathological fear of negative social evaluation, with cognitive models highlighting catastrophic anticipation of failure and compensatory safety behaviors,^17^ promoting impaired ER in social situations or during social exclusion. Individuals with SAD often over-focus on perceived performance flaws (e.g., “Does my speech seem clumsy?”) and reduce social exposure via avoidance strategies, which in turn reinforces anxiety. This cognitive-behavioral pattern not only directly interferes with the implementation of cognitive reappraisal strategies such as inhibiting self-referential processing,**^Error!^ ^Reference^ ^source^ ^not^ ^found.^**^18^ but also diminishes the effectiveness of emotion regulation by depleting prefrontal cognitive resources.^19^ At the neural level, SAD aberrant functional connectivity between ACC and DLPFC, suggesting impaired coordination between emotional appraisal and executive control networks.^20^ The co-occurrence of SAD and depression leading to a complete breakdown of the emotion regulation system, that is, depressive mood reduces social motivation, while social avoidance behaviors exacerbate social anxiety.^6^

Neuroimaging studies have highlighted the neural basis of ER, particularly via cognitive reappraisal.^21^ EEG offers real-time temporal precision, making it a clinically valuable tool for detecting and characterizing ER deficits. The late positive potential (LPP), an event-related potential beginning ∼300 ms after stimulus onset, reflects sustained attentional and affective engagement.**^Error!^ ^Reference^ ^source^ ^not^ ^found.^**^22^ Reduced LPP amplitude following cognitive reappraisal is widely interpreted as successful emotion downregulation, supported by prefrontal control over limbic reactivity.**^Error!^ ^Reference^ ^source^ ^not^ ^found.^**^23^ Additionally, frontal theta-band activity plays a critical role in cognitive control processes. Resting theta activity is associated with self-reported use of cognitive reappraisal and expressive suppression.^24^ Some studies observed an increase in theta activity when participants downregulate emotional responses to unpleasant images through reappraisal.^25^ However, contrasting findings have also shown decreased theta power under similar conditions,^26^ raising questions about the specificity and directionality of frontal theta involvement in ER. This inconsistency underscores the need for further investigation to clarify the role of frontal theta oscillations in ER among depressed adolescents. Specifically, how comorbid SAD modulates emotional regulation abilities in MDD adolescents during social exclusion, both from behavioral and EEG-based neural features or functional connectivity, is still unclear.

Using EEG in combination with a dedicated paradigm showing social exclusion (versus neutral situations), this study investigated emotion dysregulation and underlying neural mechanisms depressed adolescents with a specific focus on the moderating role of comorbid SAD. Participants were exposed to socially exclusionary stimuli while undergoing EEG recording. Behavioral indices included self-reported affect and reappraisal efficacy; neural measures focused on LPP component, frontal theta-band power spectral density, and resting-state prefrontal–limbic FC. Graph-theoretical metrics were further applied to assess network-level alterations in regulatory circuits. This multimodal approach aimed to clarify how comorbid social anxiety modulates ER capacity in depressed adolescents, both at the behavioral and neurophysiological levels. To our knowledge, this is the first study to examine both task-related and intrinsic connectivity markers of regulation impairments in this high-risk population using temporally sensitive EEG measures, providing translational insights into mechanisms of affective dysfunction and targets for early intervention.

## Materials and Methods

### Participants

A total of 62 adolescents aged 12–18 years participated were recruited, including 30 patients diagnosed with MDD from the Clinical Hospital of Chengdu Brain Science Institute, University of Electronic Science and Technology of China and 32 age-matched HCs from the community. The specific process is shown in Fig. 1A. Psychiatric diagnoses were confirmed by licensed clinicians using the structured clinical interview from the Diagnostic and Statistical Manual of Mental Disorders (DSM-V). Inclusion criteria included right-handedness, normal or corrected-to-normal vision, and a HAMD score ≥8 for the Adolescent-MDD group and <8 for HCs. Exclusion criteria were comorbid major psychiatric or neurological disorders, serious physical illness, prior neurosurgery, or receipt of neuromodulation treatments (e.g., ECT, rTMS) in the past six months. Participants completed five standardized psychological assessments on the day of the experiment, as detailed in Supplementary 1. Demographic and clinical characteristics are summarized in Table 1, there were no significant differences were observed between groups in age, gender, or education. The study was approved by the Ethics Committee of the Chengdu Fourth Hospital in China and registered at ClinicalTrials.gov (NCT06417437). The experiment was conducted in accordance with the Declaration of Helsinki. All participants provided written informed consent. Medication uses, including each anti-depressants, mood-stabilizer, antipsychotic and anxiolytic medication, were evaluated following standardized procedures, and a total medication load index was computed based on dosage coding (0 = absent, 1 = low, 2 = high).^27^ For drugs not covered above, we adopted the recommended daily dose range from the Physician’s Desk Reference, and similarly coded as 0, 1, or 2. Table 1 presents the medication load index for 30 patients; for 8 out of 30 patients medication records were not available.

**Figure 1.**
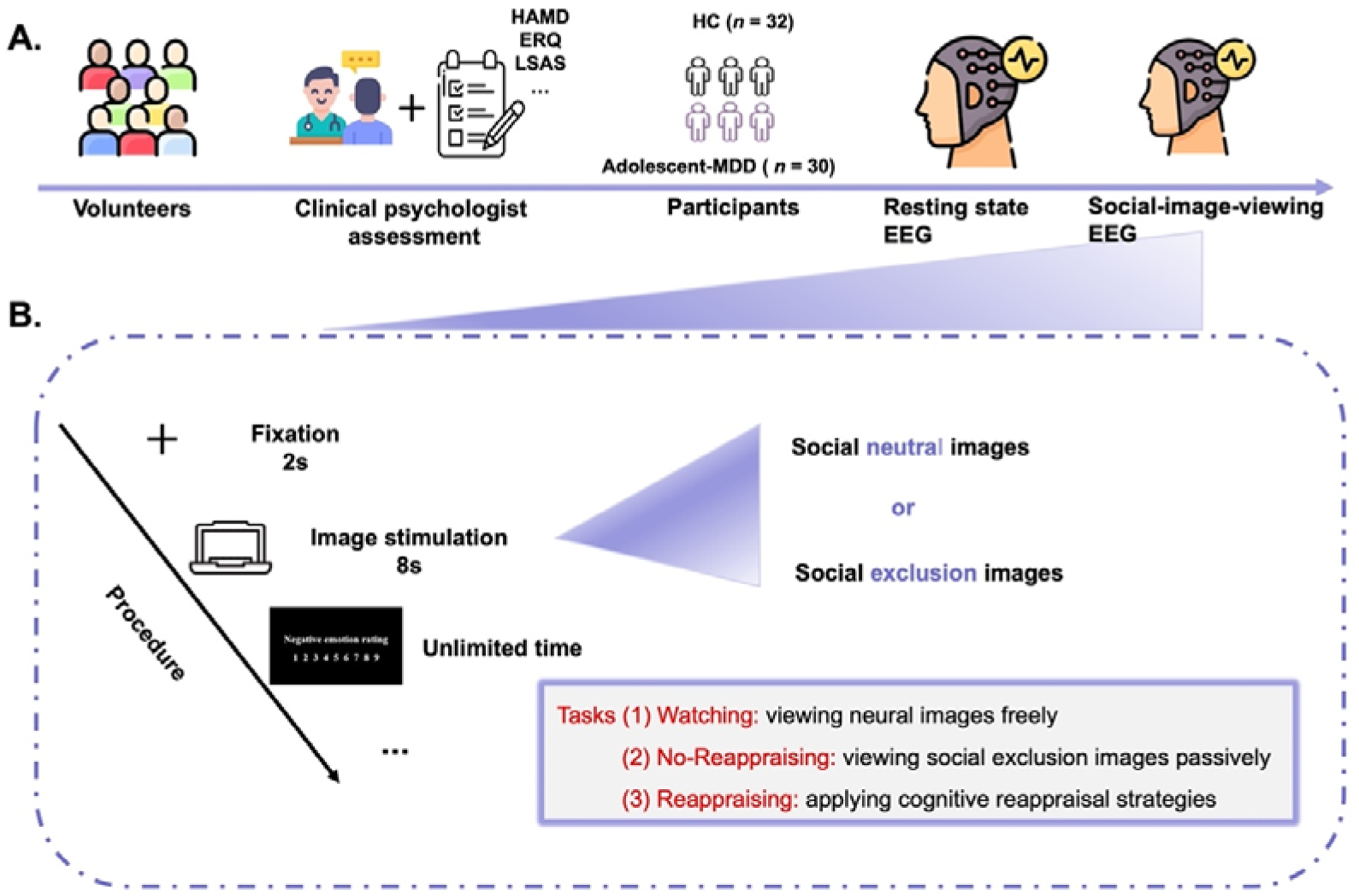
Experimental design. (A) Illustration of participants recruitment and EEG acquisition. (B) Trial structure within the social-exclusion-stimuli task. Participants were asked to conduct three conditions randomly: Watching, No-Reappraising and Reappraising.

**Table 1.**
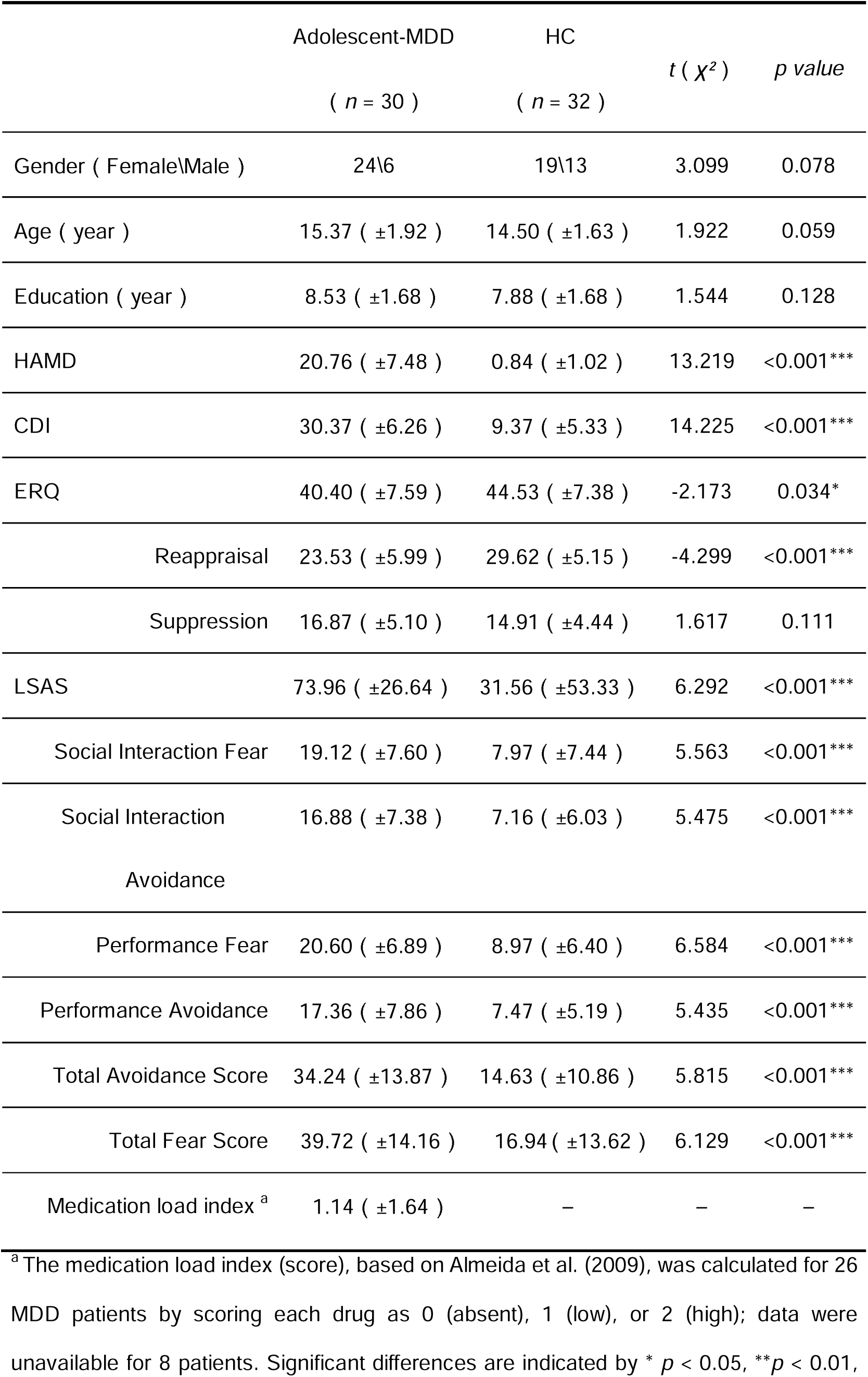
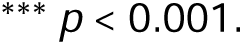
Demographics and clinical characteristics of participants (Means ± S.D.).

### Materials

All image stimuli were obtained from the Asian Youth Social Inclusion and Exclusion Image Database (ISIEA),**^Error!^ ^Reference^ ^source^ ^not^ ^found.^**^28^ a validated repository developed for Asian adolescents and publicly available through Shenzhen University, including standardized social exclusion and neutral scenarios. In the current study, 60 social exclusion images (valence: 3.43 ± 0.47), 51 social acceptance images (valence: 7.06 ± 0.20) and 53 neutral (valence: 5.11 ± 0.29) images were selected, differing significantly in valence and arousal (*F*(2,147) = 137.62, *p* < 0.001). Inter-rater reliability was high (Cronbach’s α > 0.90). Images were presented using E-Prime programming, which controls stimulus timing and order while synchronizing EEG recordings and behavioral responses to ensure consistency.

### Experimental Procedure

The experiment was conducted individually in the quiet EEG room. Each participant sat in a comfortable chair. To reduce artifacts in the EEG signal, the participant was instructed to avoid big body movements and unnecessary eye blinks.

Participants first completed a 5-minute resting-state EEG recording with eyes closed to establish baseline neural activity. After a 5-minute break, they completed the social-exclusion-stimuli task consisting of three blocks: (1) Watching condition, namely, viewing the neural images freely, (2) No-Reappraising condition, namely, viewing the social exclusion images passively, and (3) Reappraising condition, namely, applying cognitive reappraisal strategies to reduce negative affect when viewing the social exclusion images (Fig. 1B). Block order was randomized across participants to control the order effects. Each block included 30 trials. In each trial, a 2-second fixation cross was followed by an 8-second image presentation. After each trial, participants rated their negative emotional response on a 9-point scale (1 = not at all, 9 = extremely). In the No-Reappraising condition, they were asked to imagine themselves being socially excluded without attempting any cognitive regulation. In the Reappraising condition, they were instructed to reinterpret the exclusionary scene using cognitive reappraisal strategies (e.g., “I actively choose not to engage in their discussion” or “Their discussion has nothing to do with me, so I don’t need to feel excluded”) to actively alleviate their emotions. Prior to the formal data collection, participants received standardized training with 15 practice trials to ensure the correct comprehension of the tasks and the effective use of the reappraisal strategies.

### EEG Recordings and Analysis

EEG data were recorded using a 64-channel ANT Neuro system (German), with electrodes placed according to the international 10–20 system and impedance kept below 10 kΩ. Signals were sampled at 2048 Hz, referenced online to CPz, and recorded with a 0.05–100 Hz bandpass filter. An electrode below the left eye monitored ocular activity. Preprocessing was conducted in MATLAB using the EEGLAB toolbox. Data were down-sampled to 512 Hz, re-referenced to the average of M1/M2, and filtered (task-evoked EEG: 1–30 Hz; resting-state EEG: 1–40 Hz). Bad channels were interpolated, and independent component analysis (ICA) was used to remove artifacts from eye movements and muscle activity.

Event-related EEG signals were segmented into contiguous 2.5-second epochs time-locked to 500 ms pre-stimulus onset, with baseline correction applied using the 500 ms pre-stimulus window. The Late Positive Potential (LPP) was quantified by extracting the mean amplitude from electrodes P3, P4, Pz, POz, PO3, and PO4 within the 400–800 ms post-stimulus time window.^29^ Concurrently, the theta-band (4-8 Hz) neural oscillatory features were analyzed for prefrontal electrodes (F1, F3, F5, Fz, F2, F4, F6) during the identical 400–800 ms interval. For resting-state EEG, the initial and final 30 seconds of recordings were discarded, retaining a continuous 4-minute segment that was partitioned into non-overlapping 2-second epochs. Functional connectivity was computed via the Phase Locking Value (PLV) across Delta (1-4 Hz), Theta (4-8 Hz), and Alpha (8-13 Hz) frequency bands. Whole-head 60-channel PLV adjacency matrices (diagonal set to zero) were generated, followed by pointwise independent samples t-tests for group comparisons. The functional network usually contains weak connections or noise, which need to be processed through thresholding. In this paper, the topological indicators are obtained by retaining the edges that represent the connection strength of the top 40%.^30^ Statistical analyses were conducted using SPSS 27.0. To control for Type I error, all subsequently reported *p*-values from the *t*-tests were Bonferroni-corrected.

## Results

### Questionnaire Results

#### Depression and Anxiety Levels

Independent sample t-tests revealed significantly higher HAMD (*t* = 13.22, *p* < 0.001) and CDI (*t* = 14.23, *p* < 0.001) scores in the Adolescent-MDD group (Mean _HAMD_ =20.76 ±7.48) compared to HC group (Mean _HAMD_ = 0.84 ±1.02), confirming clinical levels of depression, see Table 1.

#### Emotion Regulation Ability

On the Emotion Regulation Questionnaire (ERQ), the Adolescent-MDD group reported lower cognitive reappraisal scores (*t* =-4.30, *p* < 0.001), suggesting reduced engagement in adaptive emotion regulation strategies. No group difference was observed in expressive suppression.

#### Rating of SAD

Social anxiety was measured using the Liebowitz Social Anxiety Scale (LSAS). The Adolescent-MDD group exhibited significantly higher LSAS total and subscale scores (*ts* ≥ 5.44, *ps* < 0.001), reflecting greater social anxiety and avoidance.

#### Ratings of Negative Emotions

Across all conditions, the Adolescent-MDD group reported higher negative emotion ratings than HC (*ts* ≥ 2.98, *ps* < 0.005), even under Watching conditions (*t* = 4.30, *p* < 0.001), suggesting a negative interpretation bias (Fig. 2A). Paired *t*-tests showed significantly elevated emotion ratings in the No-Reappraising vs. Watching condition (*ts* > 8.38, *ps* < 0.001), and reduced ratings in the Reappraising vs. No-Reappraising condition (*t*s > 6.11, *p*s < 0.001) for both groups. This indicates partial effectiveness of cognitive reappraisal, despite overall heightened reactivity in the Adolescent-MDD group.

**Figure 2.**
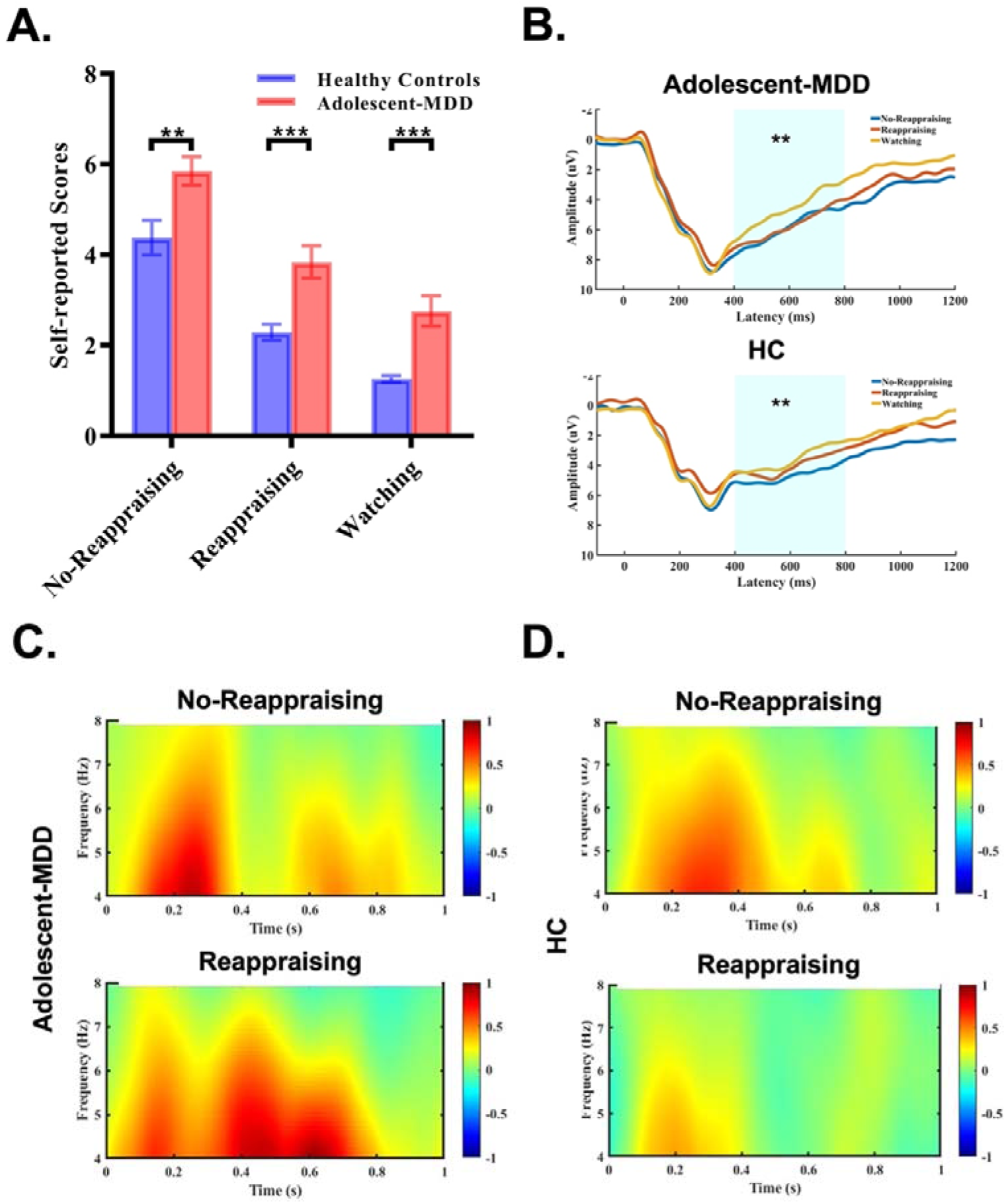
Comparison of inter-group behavioral and task-evoked neural signatures. (A) Comparison of self-reported negative emotion scores between Adolescent-MDD and HC under three experimental conditions. (B) Mean LPP amplitude across three conditions (Watching, No-Reappraising and Reappraising) for the Adolescent-MDD (top) and HC (bottom) groups. (C) and (D) Theta-band EEG oscillations in the left prefrontal cortex (F3 electrode) during the No-Reappraising/Reappraising condition in the Adolescent-MDD group and the HC group, respectively. Error bars indicate Standard Error of the Mean (SEM). Significant differences are indicated by * *p* < 0.05, ** *p* < 0.01, *** *p* < 0.001, Bonferroni corrected.

#### LPP Waveforms

A 2 (Group: MDD, HC) × 3 (Condition: Reappraise, No-Reappraise, Watch) mixed-model ANOVA was conducted on LPP amplitudes to examine the neural features of ER effects. he analysis revealed a significant main effect of group, with adolescents with MDD showing higher overall LPP amplitudes than HCs (*F*(1,60) = 5.574, *p* = 0.021), as shown in Fig. 2A. A significant main effect of condition also emerged (*F*(1,60) = 11.673, *p* = 0.001), primarily driven by lower LPP amplitudes during the Watching condition compared to the No-Reappraising condition (*t* = 2.117, *p* < 0.05; Fig. 2B); even the LPP amplitude of Reappraising remained consistent with the No-Reappraising condition (*t* = 0.855, *p* = 0.394), or the LPP amplitude of Reappraising remained consistent with the Watching condition (*t* = 1.172, *p* = 0.244). The Condition × Group interaction was not significant (*F*(2, 120) = 0.476, *p* = 0.623).

Based on the emotion regulation deficits in Adolescent-MDD, we specifically hypothesized that HCs would exhibit a significant reduction in LPP amplitude from the No-Reappraising to the Reappraising condition, whereas this modulation would be absent in the Adolescent-MDD group. To test these a priori hypotheses, planned comparisons were conducted separately for the three conditions. Those analyses revealed a critical dissociation, that is, under the Reappraising condition, Adolescent-MDD exhibited significantly greater LPP amplitudes compared to HC (*t* = 2.419, *p* = 0.019). Under the No-Reappraising condition, Adolescent-MDD exhibited marginally greater LPP amplitudes compared to HC (*t* = 1.929, *p* = 0.058), and under the Watching condition, both groups exhibited non-significant the modulation patterns of LPP amplitudes (*t* = 1.905, *p* = 0.062). Within-group comparisons further indicated that HC exhibited a significant LPP reduction from maintenance to regulation (*t* = 3.038, *p* = 0.005), indicating effective downregulation of emotional responses. In contrast, Adolescent-MDD showed no significant reduction (*t* = 0.599, *p* = 0.554), suggesting impaired physiological regulation.

### Altered Task-Related and Resting-State Neural Oscillations in Depressed Adolescents

Repeated-measures ANOVAs examined the neural oscillations of ER effects on theta-band in HC and MDD groups. As shown in Fig. 2C&D, the Adolescent-MDD group showed significantly higher overall theta-band activities than the HC group (*F*(1,60) = 1.851, *p* = 0.179). Although the main effects of Condition and Group were not significant (Condition: *F*(1,60) = 0.869, *p* = 0.422, Group: *F*(1,60) = 1.851, *p* = 0.179), the Condition × Group interaction was significant (*F*(2, 120) = 4.884, *p* < 0.05). Notably, post-hoc analyses revealed a significant group difference in the left anterior prefrontal cortex, specifically, the Adolescent-MDD group exhibited significantly greater theta power than the HC group at the F3 electrode site (*t* = 2.542, *p* = 0.014; Fig. 2C&D), which was part of the targeted F1, F3, and F5 electrode cluster during the Reappraising condition. This result indicated that Adolescent-MDD need more additional resources when applying cognitive reappraisal strategies.

For resting-state EEG oscillations, time–frequency analysis revealed significantly higher low-frequency (1–13 Hz) EEG power in Adolescent-MDD than that in HC group, particularly in the frontal region. Notably, Delta (1–4 Hz, *t* = 2.550, *p* = 0.012), Theta (4–8 Hz, *t* = 4.081, *p* < 0.001), and Alpha (8–13 Hz, *t* = 3.310, *p* = 0.002) bands were significantly increased in the Adolescent-MDD group, see Table 2.

**Table 2.** Comparison of different frequency bands in the prefrontal cortex between adolescents with Adolescent-MDD and HC group.

| | Adolescent-MDD ($\mu V^2$) | HC ($\mu V^2$) | $t$ | $p$ |
| --- | --- | --- | --- | --- |
| Delta-band<br>(1-4 Hz) | 21.97 ($\pm 16.80$) | 13.23 ($\pm 8.64$) | 2.55 | 0.012* |
| Theta-band<br>(4-8 Hz) | 7.63 ($\pm 4.86$) | 3.83 ($\pm 2.01$) | 4.081 | <0.001*** |
| Alpha-band<br>(8-13 Hz) | 9.30 ($\pm 5.98$) | 5.05 ($\pm 3.99$) | 3.310 | 0.002** |

Adolescent-MDD: Adolescents with Major Depressive Disorder; HC: Healthy Control.
Significant differences are indicated by \* $p < 0.05$ , \*\* $p < 0.01$ , \*\*\* $p < 0.001$ .

### Multi-dimensional Associations Among Task-related Change in Neural, Clinical, and Behavioral Measures

To explore the neural mechanism of emotional regulation for Adolescent-MDD, multi-dimensional association analysis was conducted among EEG, clinical and cognitive measures.

At the behavioral level, HAMD and CDI scores separately showed positive correlations with higher negative emotion ratings across multiple conditions of the social-exclusion stimuli task (*rs* > 0.323, *ps* < 0.014). Notably, the factor of performance avoidance in the LSAS scores was significantly correlated with the Reappraising condition (*r* = 0.491, *p* < 0.001), rather than No-Reappraising condition (*r* = 0.253, *p* = 0.058), suggesting that socially anxious individuals may underreport negative affect due to emotional avoidance. For details, see Supplementary Table S1 and Table S2. These findings question the reliability of self-report ratings as objective indices of regulation success in this population.

From all participants, LPP amplitude of Reappraising condition was positively correlated with HAMD, CDI, and LSAS scores (*rs* > 0.275, *ps* < 0.038, see Table S3), indicating that greater neural reactivity during attempted emotional regulation was associated with more severe depressive and social anxiety symptoms. Furthermore, the theta-band power in the left prefrontal brain region was positively correlated with HAMD, HAMA, and LSAS scores (*r*s > 0.270, *p*s < 0.034, see Table S4), suggesting that higher emotional symptom severity may be associated with increased compensatory recruitment of prefrontal cognitive control resources during emotion regulation in depression. Significant correlations were also found in the three frequency bands of the resting state with HAMD, CDI and LSAS scores (*rs* > 0.315, *ps* < 0.017, see Table S5), suggesting that increased resting-state low-frequency activity may reflect mood and anxiety symptom severity.

### Altered Resting-State Network Dynamics in Adolescent Depression

We examined group connectivity differences in PLV analysis to reveal altered resting-state FC in depressed adolescents, details shown in Table 3. Shown as the Fig. 3A, compared to HCs, the Adolescent-MDD group showed significantly increased connectivity in the left frontal region in the delta-band connections (*t* = 2.550, *p* = 0.012), and stronger frontal-central and interhemispheric connections in the theta-band (*t* = 3.566, *p* < 0.001), see Fig. 3B. Shown as the Fig. 3C, alpha-band connections were also elevated in the left frontal lobe (*t* = 3.310, *p* = 0.002) for the Adolescent-MDD group. Shown as Table 3, topological analysis further showed reduced global efficiency in the Adolescent-MDD group across delta, theta, and alpha bands (*ts* <-2.115, *ps* < 0.039), with the largest effect in the theta-band (*t* =-2.662, *p* = 0.010). Average path length was longer in the Adolescent-MDD group than the HC group across three frequency bands (*ts* > 1.806, *ps* < 0.041). Similarly, in the Adolescent-MDD group, modularity and clustering coefficient were significantly higher in the delta and theta bands (*ts* > 2.435, *ps* < 0.018), indicating impaired integration but enhanced local segregation in brain networks.

**Figure 3.**
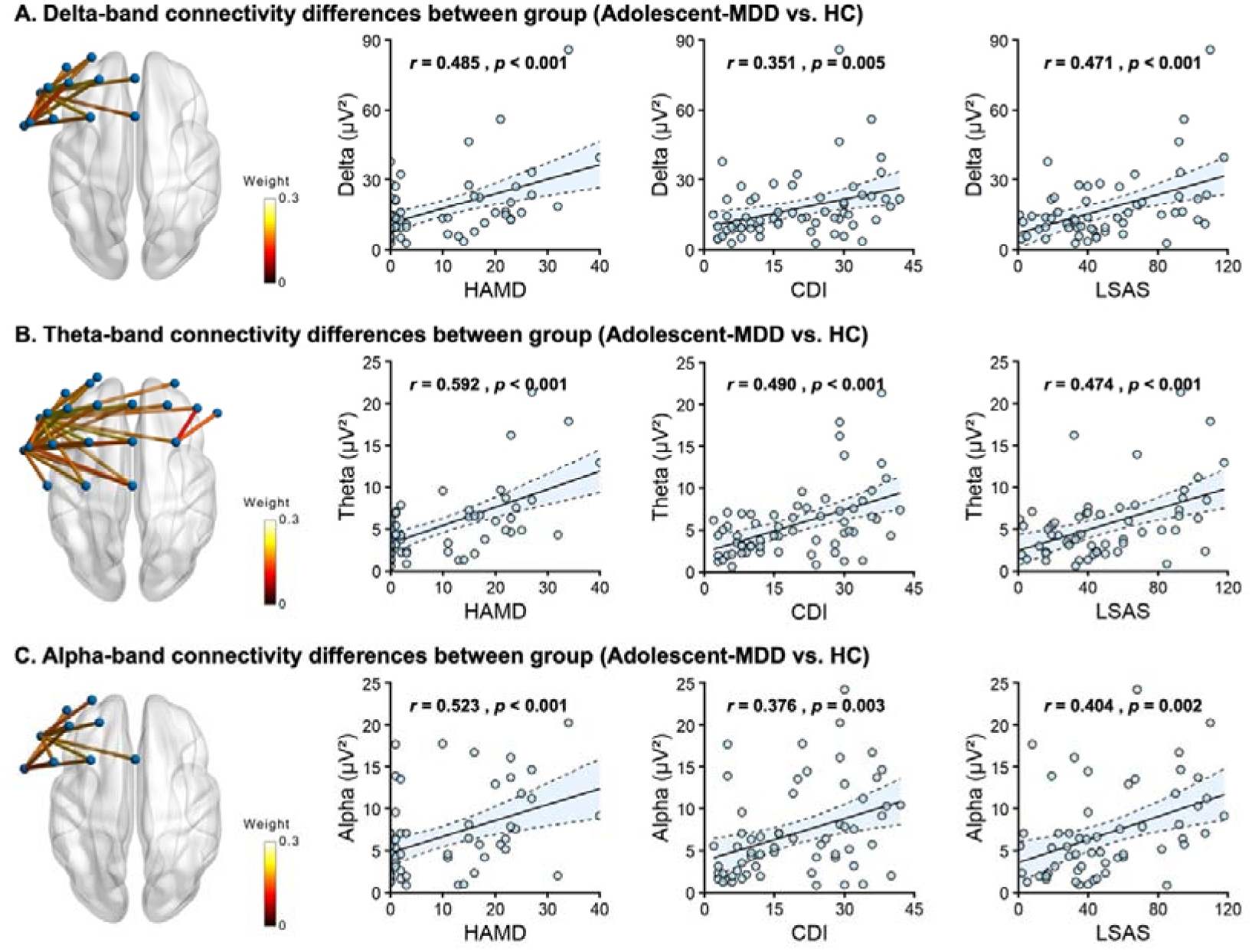
Group differences in functional connectivity as measured by PLV and correlations with clinical symptoms. The left panels in each sub-plots (A-C) illustrate significant PLV differences between the Adolescent-MDD and HC groups, calculated by subtracting HC group values from MDD group values to highlight the points of significant connectivity differences. These group differences are displayed for the (A) delta-band power, (B) theta-band power, and (C) alpha-band, respectively. Additionally, the right panels in (A-C) represents the correlations between PLV difference in the delta, theta, and alpha bands and scores on the HAMD, CDI, and LSAS, respectively. The color bar indicates the magnitude of the PLV difference, while L and R denote the left and right hemispheres.

**Table 3.**
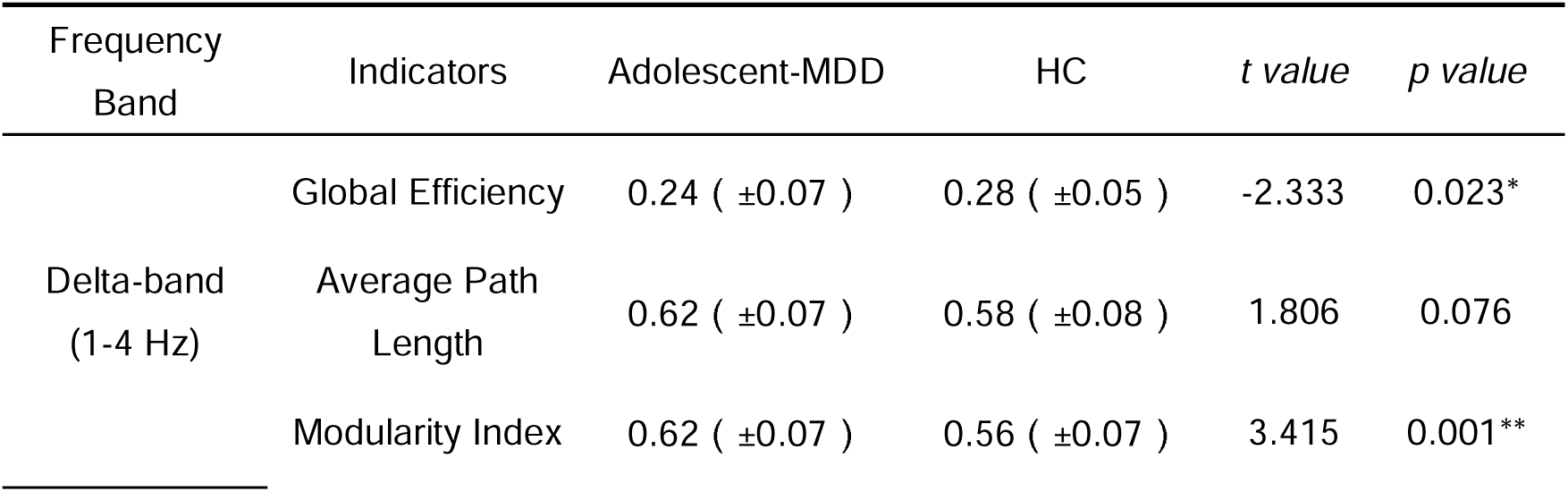
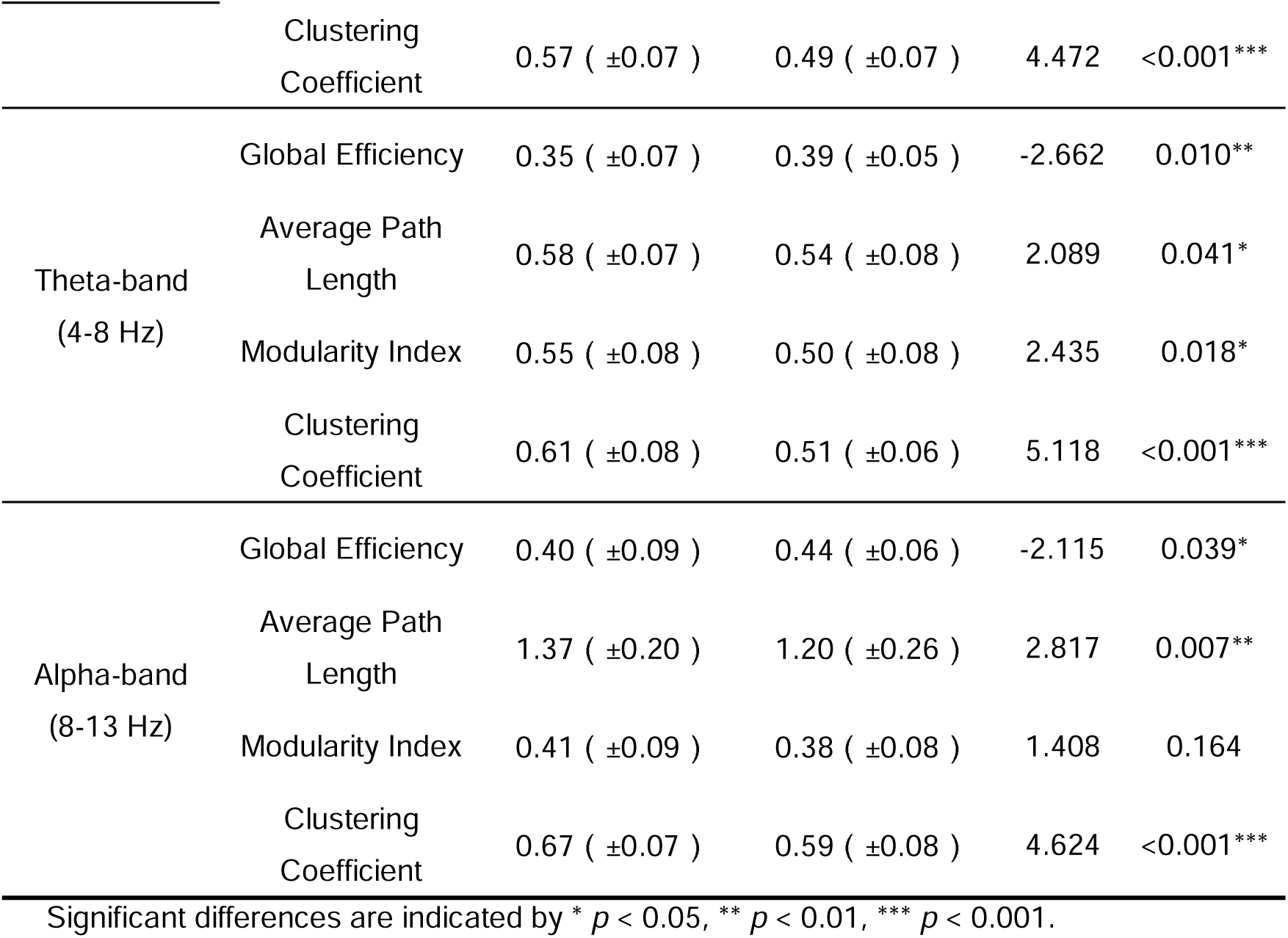
EEG Indicators for Different Frequency Bands in Adolescent-MDD and HC Groups.

## Discussion

This study aims to investigate neurophysiological correlates underlying emotion dysregulation in depressed adolescents during observation of social exclusion, with a specific focus on how social anxiety symptoms shape cognitive reappraisal processes. We assessed behavioral, electrophysiological, and brain connectivity-based markers in an Adolescent-MDD and a HC group under three emotional induced conditions. We found that adolescents with MDD exhibited pronounced impairments in emotion regulation, as indicated by lower self-reported use of cognitive reappraisal strategies and stronger negative affect across all task conditions. Notably, greater depressive severity was associated with heightened social anxiety, which further attenuated the ability to downregulate negative affect during social exclusion. These findings suggest that social anxiety is not only comorbid with depression but may actively interfere with the use of regulatory strategies in socially stressful contexts.

At the neurophysiological level, depressed adolescents showed increased LPP amplitudes and higher theta-band activity in the left prefrontal cortex during cognitive reappraisal processing. These results align with existing literature, indicating that depressed individuals often exhibit abnormal neurophysiological activity during emotional processing. For instance, Hajcak et al. (2009)^31^ pointed out that depressed patients showed abnormal LPP responses during emotional processing, especially when handling negative emotional stimuli, with their emotional regulation abilities often impaired. Similarly, theta-band power increases during regulation may reflect compensatory recruitment of prefrontal resources in response to heightened emotional challenge.^32^ In the current study, we observed a significant positive correlation between the increase in theta frequency band power spectral density during the Reappraising condition and the severity of SAD (i.e., LSAS score) in the left prefrontal region. This suggests that social anxiety may play a critical role in emotional regulation. The functional compensation theory posits that when an individual’s emotional regulation capacity is limited, they rely on additional neurophysiological efforts to maintain relatively normal emotional responses.^33^ This effort often manifests as increased neural activity, as observed in our study. When depressed adolescents face social situations, excessive anxiety may lead to more pronounced functional compensation in the prefrontal cortex. In other words, SAD may be a key factor contributing to the additional consumption of neural resources in the emotional regulation process when confronted with social scenarios.

Resting-state results that depressed adolescents exhibited significant enhancement of low-frequency power in the delta, theta, and alpha frequency-bands at resting state, particularly in the frontal lobe. These patterns are consistent with neuroimaging evidence from previous studies on adolescent depression, suggesting that the prefrontal cortex may be structurally or functionally immature.^34^ The prefrontal cortex is a core brain region for emotion regulation, and its myelination process is not completed during adolescence,^35^ leading to lower efficiency in processing emotional information. The enhancement of low-frequency EEG activity may reflect abnormal connections between the prefrontal lobe and the limbic system (such as the amygdala), manifesting as a disinhibited neural network state at rest.^36^

Specifically, the abnormality in the delta frequency band may be related to overactivation of the default mode network, which weakens the ability of prefrontal cortex to integrate autobiographical memory and exacerbates emotional rumination**^Error!^ ^Reference^ ^source^ ^not^ ^found.^**.^11^ Meanwhile, the increase in the theta-band frequency band may reflect impaired regulation by the anterior cingulate-dorsolateral prefrontal pathway, which is responsible for inhibiting irrelevant emotional stimuli. Dysfunction in this pathway leads to excessive occupation of attention resources by negative information.^37^ Notably, the increase in alpha-band activity in the frontal lobe of the Adolescent-MDD group was significantly positively correlated with SAD. Alpha oscillations are typically associated with inhibitory control of cognitive resources, and their enhancement may indicate that depressed adolescents are more likely to adopt passive avoidance strategies rather than actively regulate emotions during the resting state.^38^ This finding is consistent with behavioral studies indicating that SAD exacerbates emotional dysregulation, suggesting that abnormalities in prefrontal resting-state activity may underlie the co-occurrence of social anxiety and depressive symptoms.

Resting-state FC results revealed enhanced synchronization within the left prefrontal cortex in the Adolescent-MDD group, but a reduction in global efficiency. The result is consistent with the fMRI findings by a similar cognitive reappraisal condition for depressed adolescents, which demonstrated reduced connectivity between the left dorsomedial prefrontal cortex (DMPFC) and the anterior insula/inferior frontal gyri bilaterally (AI/IFG) and between left DLPFC and left AI/IFG.^39^ This pattern of increased local connectivity and decreased global integration may be related to delayed development of the anterior cingulate-dorsolateral prefrontal network in adolescents with depression. The anterior cingulate is involved in emotional conflict monitoring, while the dorsolateral prefrontal cortex is responsible for cognitive control. A lack of coordination between these regions leads to decreased emotion regulation efficiency.^40^ Given adolescent neurodevelopmental characteristics, the delay in this network may be linked to delayed myelination^41^ or changes in dopamine receptor sensitivity,^42^ which impair the prefrontal cortex’s ability to integrate emotional signals from the limbic system. Furthermore, the reduced global efficiency observed in the delta frequency-band suggests that interregional brain communication is less effective in depressed adolescents at rest. This inefficiency may be attributable to slower nerve conduction resulting from abnormal synaptic plasticity or insufficient myelination.^43^ Such a decline in network efficiency could, in turn, weaken the prefrontal cortex’s regulation of the limbic system. This process may establish a self-perpetuating loop where an enhanced emotional response compromises prefrontal control, thereby exacerbating subsequent emotional dysregulation.**^Error!^ ^Reference^ ^source^ ^not^ ^found.^**^44^

Abnormalities in resting low-frequency activity provide potential biomarkers for early identification of adolescent depression and may guide treatment development and evaluation. For example, increased theta power in the frontal lobe and abnormal anterior cingulate-dorsolateral prefrontal connectivity have been associated with the effectiveness of cognitive behavioral therapy (CBT).^45^ CBT improves abnormal theta-band activity by enhancing the prefrontal cortex’s ability to cognitively reappraise emotional cues. Our findings suggest that future interventions targeting the maturation of the prefrontal neural network could include training programs designed to promote myelination and functional integration, such as transcranial magnetic stimulation (TMS) or neurofeedback. TMS can accelerate myelination by modulating prefrontal neuron excitability,^46^ while neurofeedback may help patients actively inhibit the overactive default mode network by providing real-time feedback on alpha-band activity^47^ or to gain better regulatory control over prefrontal executive functions (e.g. Yang et al. 2024; Li et al., 2019),^48,49^ thereby improving emotion regulation abilities.

In conclusion, our findings identify a distinct neurophysiological profile of emotion dysregulation in depressed adolescents, characterized by enhanced low-frequency resting-state activity, disrupted global network efficiency, and heightened task-related prefrontal responses during cognitive reappraisal condition. These abnormalities were most pronounced in individuals with greater social anxiety symptoms, suggesting that anxiety-related mechanisms may exacerbate neural inefficiency in affective control networks. The convergence of resting-state and task-evoked indices points to shared disruptions in prefrontal-limbic circuitry underlying both spontaneous and effortful emotion regulation. These results extend prior work by highlighting the moderating role of social anxiety in shaping compensatory neural responses to emotional demands. Future studies should examine whether these electrophysiological signatures—particularly increased frontal theta activity and reduced network integration—can serve as biomarkers to predict treatment response or guide the development of neuro-modulatory interventions aimed at enhancing cognitive control in adolescents with comorbid depression and social anxiety.

## Code & Data Availability

The codes and data supporting the findings of this study are available from the corresponding author upon reasonable request and with permission from the university administration.

## Acknowledgments

This work was funded by Science and Technology Innovation (STI) 2030-Major Projects (No. 2022ZD0208500); Sichuan Province Postdoctoral Research Special Funding Project (No. TB2025066); National Natural Science Foundation of China (No.62276051).

## Competing Interests

The authors declare no competing interests.

## Notes

### Competing Interest Statement

The authors have declared no competing interest.

### Clinical Trial

NCT06417437

### Author Declarations

Ethics Committee of the Chengdu Fourth Hospital in China gave ethical approval for this work.

