## Supplemental Table1to5 for "Prefrontal-Limbic Dysconnectivity Underlies Emotion Regulation Deficits in Depressed Adolescents with Anxiety Disorder"

**Supplementary**

**Supplement 1. Introduction to the Psychological Measurement Tools Used in the Study.**

The Hamilton Depression Rating Scale (HAMD)^[1]^ is used to assess the severity of depression and consists of 17 items covering symptoms such as mood, sleep, appetite, energy, and anxiety. Each item is scored on a scale from 0 to 4, where 0 indicates no symptoms and 4 indicates the most severe symptoms. A higher total score reflects more severe depressive symptoms. HAMD has demonstrated high internal consistency (Cronbach's α = 0.88) and has been validated through multiple studies, effectively distinguishing different levels of depression severity.

The Children's Depression Inventory (CDI)^[2]^ is designed to assess the severity of depressive symptoms in children and adolescents. The scale includes 27 items, addressing mood, behavior, and cognition, such as sadness, loneliness, and negative views about the future. Each item is scored on a scale from 0 to 2 based on the frequency of symptoms, with higher scores indicating more severe depression. CDI is widely used for screening and assessing depressive symptoms in children and adolescents, with strong internal consistency (Cronbach’s α = 0.86) and good construct validity.

The Hamilton Anxiety Rating Scale (HAMA)^[3]^ is used to evaluate the severity of anxiety disorders, particularly generalized anxiety disorder and social anxiety disorder. The scale contains 14 items that assess physical symptoms, psychological symptoms, and behavioral manifestations of anxiety. Each item is rated on a scale of 0 to 4, where 0 indicates no symptoms and 4 indicates the most severe symptoms. Higher total scores reflect greater anxiety severity. HAMA has excellent internal consistency (Cronbach’s α = 0.93) and is widely used in both clinical practice and research.

The Emotion Regulation Questionnaire (ERQ)^[4]^ assesses two main strategies used in emotion regulation: cognitive reappraisal and emotion suppression. The scale consists of 10 items, with 6 items related to cognitive reappraisal and 4 related to emotion suppression. Each item is scored on a 7-point Likert scale, where higher scores indicate a greater tendency to use a particular emotion regulation strategy. ERQ shows high internal consistency (Cronbach's α = 0.85) and strong construct validity, making it a widely used tool for examining emotion regulation strategies and their impact on mental health.

The Liebowitz Social Anxiety Scale (LSAS)^[5]^ is designed to assess the severity of social anxiety disorder. The scale includes 24 items, divided into two dimensions: anxiety and avoidance in social situations. Each item is rated from 0 to 3, with 0 indicating no anxiety or avoidance, and 3 indicating extreme anxiety or avoidance. The total LSAS score reflects the severity of social anxiety symptoms, with higher scores indicating more severe symptoms. LSAS has strong internal consistency (Cronbach’s α = 0.93) and excellent test-retest reliability (r = 0.88), making it a widely used tool for diagnosing and evaluating the treatment of social anxiety disorder.

**Supplement 2. Tables to Dimensional Associations Across Cognitive, EEG, and Clinical Measures.**

**Table S1. Correlations between the Emotional Scores of All Participants and** **Clinical Symptom Severity.**

|  | Watch | | No-Reappraising | | Reappraising | |
| --- | --- | --- | --- | --- | --- | --- |
|  | *r* | *p* | *r* | *p* | *r* | *p* |
| HAMD | 0.607** | < 0.001 | 0.323* | 0.014 | 0.531** | <0.001 |
| CDI | 0.639** | < 0.001 | 0.354** | 0.005 | 0.528** | <0.001 |
| ERQ | -0.072 | 0.578 | <0.001 | 0.998 | 0.010 | 0.938 |
| Reappraisal | -.251* | 0.049 | -0.072 | 0.579 | -0.104 | 0.420 |
| Suppression | 0.214 | 0.095 | 0.093 | 0.470 | 0.153 | 0.237 |
| LSAS | 0.477** | <0.001 | 0.229 | 0.086 | 0.439** | 0.001 |
| Social Interaction Fear | 0.394** | 0.002 | 0.203 | 0.13 | 0.384** | 0.003 |
| Social Interaction Avoidance | 0.502** | <0.001 | 0.235 | 0.079 | 0.353** | 0.007 |
| Performance Fear | 0.377** | 0.004 | 0.180 | 0.180 | 0.435** | 0.001 |
| Performance Avoidance | 0.542** | <0.001 | 0.253 | 0.058 | 0.491** | <0.001 |
| Total Avoidance Score | 0.545** | <0.001 | 0.255 | 0.056 | 0.440** | 0.001 |
| Total Fear Score | 0.391** | 0.003 | 0.194 | 0.148 | 0.414** | 0.001 |

Significant differences are indicated by * *p* < 0.05, ** *p* < 0.01, *** *p* < 0.001.

**Table S2. Correlations between Reappraising Scores and Clinical Symptom Severity.**

|  | Reappraising | | | |
| --- | --- | --- | --- | --- |
|  | Adolescent-MDD | | HC | |
|  | *r* | *p* | *r* | *p* |
| HAMD | 0.399* | 0.048 | 0.126 | 0.491 |
| CDI | 0.494** | 0.006 | -0.084 | 0.649 |
| ERQ | 0.137 | 0.471 | 0.210 | 0.250 |
| Reappraisal | 0.138 | 0.466 | 0.188 | 0.302 |
| Suppression | 0.041 | 0.830 | 0.130 | 0.480 |
| LSAS | 0.268 | 0.195 | 0.060 | 0.743 |
| Social Interaction Fear | 0.166 | 0.428 | 0.088 | 0.631 |
| Social Interaction Avoidance | 0.159 | 0.448 | -0.018 | 0.920 |
| Performance Fear | 0.224 | 0.282 | 0.101 | 0.582 |
| Performance Avoidance | 0.401* | 0.047 | 0.041 | 0.822 |
| Total Avoidance Score | 0.312 | 0.129 | 0.010 | 0.958 |
| Total Fear Score | 0.198 | 0.343 | 0.096 | 0.602 |

Significant differences are indicated by * *p* < 0.05, ** *p* < 0.01, *** *p* < 0.001.

**Table S3. Correlations between Reappraise-Condition LPP Amplitude and Clinical Scale Scores.**

|  | ALL | | Adolescent-MDD | | HC | |
| --- | --- | --- | --- | --- | --- | --- |
|  | *r* | *p* | *r* | *p* | *r* | *p* |
| HAMD | 0.277* | 0.037 | 0.114 | 0.588 | -0.037 | 0.84 |
| CDI | 0.329** | 0.009 | 0.390* | 0.033 | -0.141 | 0.44 |
| LSAS | 0.299* | 0.024 | 0.307 | 0.136 | -0.032 | 0.864 |
| Social Interaction Fear | 0.313* | 0.018 | 0.386 | 0.057 | -0.024 | 0.897 |
| Social Interaction Avoidance | 0.293* | 0.027 | 0.282 | 0.172 | -0.003 | 0.989 |
| Performance Fear | 0.286* | 0.031 | 0.29 | 0.16 | -0.047 | 0.797 |
| Performance Avoidance | 0.234 | 0.08 | 0.148 | 0.481 | -0.046 | 0.801 |
| Total Avoidance Score | 0.275* | 0.038 | 0.234 | 0.261 | -0.024 | 0.898 |
| Total Fear Score | 0.304* | 0.022 | 0.348 | 0.088 | -0.035 | 0.848 |

ALL: All participants. Significant differences are indicated by * *p* < 0.05, ** *p* < 0.01, *** *p* < 0.001.

**Table S4. Correlations of Left Prefrontal Theta-band Power Density with Clinical Scores During the Social-exclusion-stimuli Task.**

|  | Watch | | No-Reappraising | | Reappraising | |
| --- | --- | --- | --- | --- | --- | --- |
|  | *r* | *p* | *r* | *p* | *r* | *p* |
| HAMD | -0.223 | 0.095 | 0.071 | 0.601 | 0.374** | 0.004 |
| CDI | -0.156 | 0.227 | 0.005 | 0.966 | 0.270* | 0.034 |
| LSAS | -0.225 | 0.092 | 0.053 | 0.693 | 0.338** | 0.01 |
| Social Interaction Fear | -0.259 | 0.052 | 0.021 | 0.88 | 0.261 | 0.050 |
| Social Interaction Avoidance | -0.122 | 0.366 | 0.148 | 0.272 | 0.334** | 0.011 |
| Performance Fear | -0.258 | 0.053 | -0.005 | 0.972 | 0.319* | 0.015 |
| Performance Avoidance | -0.203 | 0.13 | 0.046 | 0.732 | 0.370** | 0.005 |
| Total Avoidance Score | -0.17 | 0.207 | 0.102 | 0.452 | 0.367** | 0.005 |
| Total Fear Score | -0.261* | 0.049 | 0.008 | 0.951 | 0.293* | 0.027 |

Significant differences are indicated by * *p* < 0.05, ** *p* < 0.01, *** *p* < 0.001.

**Table S5. Correlations Between Resting-State Frontal Lobe Theta Power and Clinical Scores Across All Participants.**

|  | ALL | | | | | | |
| --- | --- | --- | --- | --- | --- | --- | --- |
|  | Delta-band  (1-4 Hz) | | Theta-band  (4-8 Hz) | | | Alpha-band  (8-13Hz) | |
|  | *r* | *p* | *r* | *p* | | *r* | *p* |
| HAMD | 0.485*** | <0.001 | 0.592*** | <0.001 | | 0.523*** | <0.001 |
| CDI | 0.351** | 0.005 | 0.490*** | <0.001 | | 0.376** | 0.003 |
| ERQ | -0.165 | 0.201 | -0.282* | 0.026 | | -0.113 | 0.383 |
| Reappraisal | -0.276* | 0.03 | -0.348** | | 0.006 | -0.173 | 0.18 |
| Suppression | 0.099 | 0.444 | 0.006 | 0.964 | | 0.046 | 0.722 |
| LSAS | 0.471*** | <0.001 | 0.474*** | <0.001 | | 0.404** | 0.002 |
| Social Interaction Fear | 0.407** | 0.002 | 0.420*** | 0.001 | | 0.315* | 0.017 |
| Social Interaction Avoidance | 0.424*** | 0.001 | 0.431*** | 0.001 | | 0.439*** | 0.001 |
| Performance Fear | 0.484*** | <0.001 | 0.478*** | <0.001 | | 0.338** | 0.01 |
| Performance Avoidance | 0.467*** | <0.001 | 0.464*** | <0.001 | | 0.450*** | <0.001 |
| Total Avoidance Score | 0.465*** | <0.001 | 0.467*** | <0.001 | | 0.464*** | <0.001 |
| Total Fear Score | 0.450*** | <0.001 | 0.453*** | <0.001 | | 0.330** | 0.012 |

ALL: All participants. Significant differences are indicated by * *p* < 0.05, ** *p* < 0.01, *** *p* < 0.001.
